# Unsupervised Machine Learning Unveil Easily Identifiable Subphenotypes of COVID-19 With Differing Disease Trajectories

**DOI:** 10.1101/2023.04.07.23288152

**Authors:** Jacky Chen, Jocelyn Hsu, Alexandra Szewc, Clotilde Balucini, Tej D. Azad, Kirby Gong, Han Kim, Robert D Stevens

## Abstract

**Background:** Given the clinical heterogeneity of COVID-19 infection, we hypothesize the existence of subphenotypes based on early inflammatory responses that are associated with mortality and additional complications.

**Methods:** For this cross-sectional study, we extracted electronic health data from adults hospitalized patients between March 1, 2020 and May 5, 2021, with confirmed primary diagnosis of COVID-19 across five Johns Hopkins Hospitals. We obtained all electronic health records from the first 24h of the patient’s hospitalization. Mortality was the primary endpoint explored while myocardial infarction (MI), pulmonary embolism (PE), deep vein thrombosis (DVT), stroke, delirium, length of stay (LOS), ICU admission and intubation status were secondary outcomes of interest. First, we employed clustering analysis to identify COVID-19 subphenotypes on admission with only biomarker data and assigned each patient to a subphenotype. We then performed Chi-Squared and Mann-Whitney-U tests to examine associations between COVID-19 subphenotype assignment and outcomes. In addition, correlations between subphenotype and pre-existing comorbidities were measured using Chi-Squared analysis.

**Results:** A total of 7076 patients were included. Analysis revealed three distinct subgroups by level of inflammation: hypoinflammatory, intermediate, and hyperinflammatory subphenotypes. More than 25% of patients in the hyperinflammatory subphenotype died compared to less than 3% hypoinflammatory subphenotype (p<0.05). Additional analysis found statistically significant increases in the rate of MI, DVT, PE, stroke, delirium and ICU admission as well as LOS in the hyperinflammatory subphenotype.

**Conclusion:** We identify three distinct inflammatory subphenotypes that predict a range of outcomes, including mortality, MI, DVT, PE, stroke, delirium, ICU admission and LOS. The three subphenotypes are easily identifiable and may aid in clinical decision making.

## Introduction

Since the first coronavirus disease (COVID-19) case reported in December 2019 (1), it was clear that this disease has extraordinarily heterogenous patient presentations that lead to differing outcomes. The heterogeneity in the patient presentation makes it difficult for clinicians to predict which patients are likely to deteriorate, resulting in delayed treatment. A clearer understanding of patient heterogeneity and disease trajectory might help clinicians anticipate which patients are likely to require more aggressive and earlier treatments and which therapies will be most beneficial to a patient.

One of the factors associated with worse outcomes is the presence of an overactive immune system such as a cytokine storm, a hyperinflammatory state secondary to excessive cytokine production (2,3). Additionally, research has shown that different types of immune-responses are associated with differing levels of disease severity (4). Although resolving this heterogeneity would allow for a more individualized treatment approach, it remains difficult to do so only using standard clinical features early on in the disease trajectory.

Our primary objective was to discover distinct subphenotypes of COVID-19 that would be easily identifiable by a human practitioner early in the disease trajectory, providing order and structure to the heterogeneity mentioned above. Although there already exists limited literature on developing subphenotypes of COVID-19, previous studies either have low sample sizes, generate subphenotypes that are difficult to distinguish by clinicians, or are hard to understand in the context of immune responses (5–8). Our study aims to overcome these limitations and produce subphenotypes that can aid timely clinical decision-making and lead to better patient outcomes.

## Methods

### Data

The data utilized for this study was taken from the JH-CROWN: The COVID PMAP Registry, which contains data from five Johns Hopkins Hospitals serving approximately 7 million people in the Maryland/Washington D.C. area and containing over 2,000 beds (9–12). The database was constructed directly from the clinical electronic health records and contains hospitalized adult patients (>18y) who were diagnosed with COVID-19 between March 5, 2020 and May 5, 2021. Diagnosis of COVID-19 was defined as the following: the detection of SARS-CoV-2 from any nucleic acid test of any specimen type with an Emergency Use Authorization from the U.S., patients who had been flagged as having COVID-19, or who had been diagnosed with suspected or confirmed COVID-19 at discharge (13).

### Patient Selection

Within our study’s timeframe, there were 15,470 COVID-19 hospital stays from the JH-CROWN database that includes hospitalized patients with any COVID-19 diagnosis. We excluded patients who were hospitalized for other ailments but had COVID-19 as a secondary diagnosis (n = 8378). For example, a patient who was hospitalized for trauma following a car accident and contracted COVID-19 while hospitalized would be excluded. Then, we excluded patients who have no recorded laboratory tests within the first 24 hours of admission (n = 16), leaving 7076 patients to be included in our study. To summarize, the 7076 patients examined in this study have primary diagnosis of COVID 19 and have recorded laboratory tests within the first 24 hours of admission. Figure 1 contains a graphical representation of the patient selection process.

**Figure 1:**
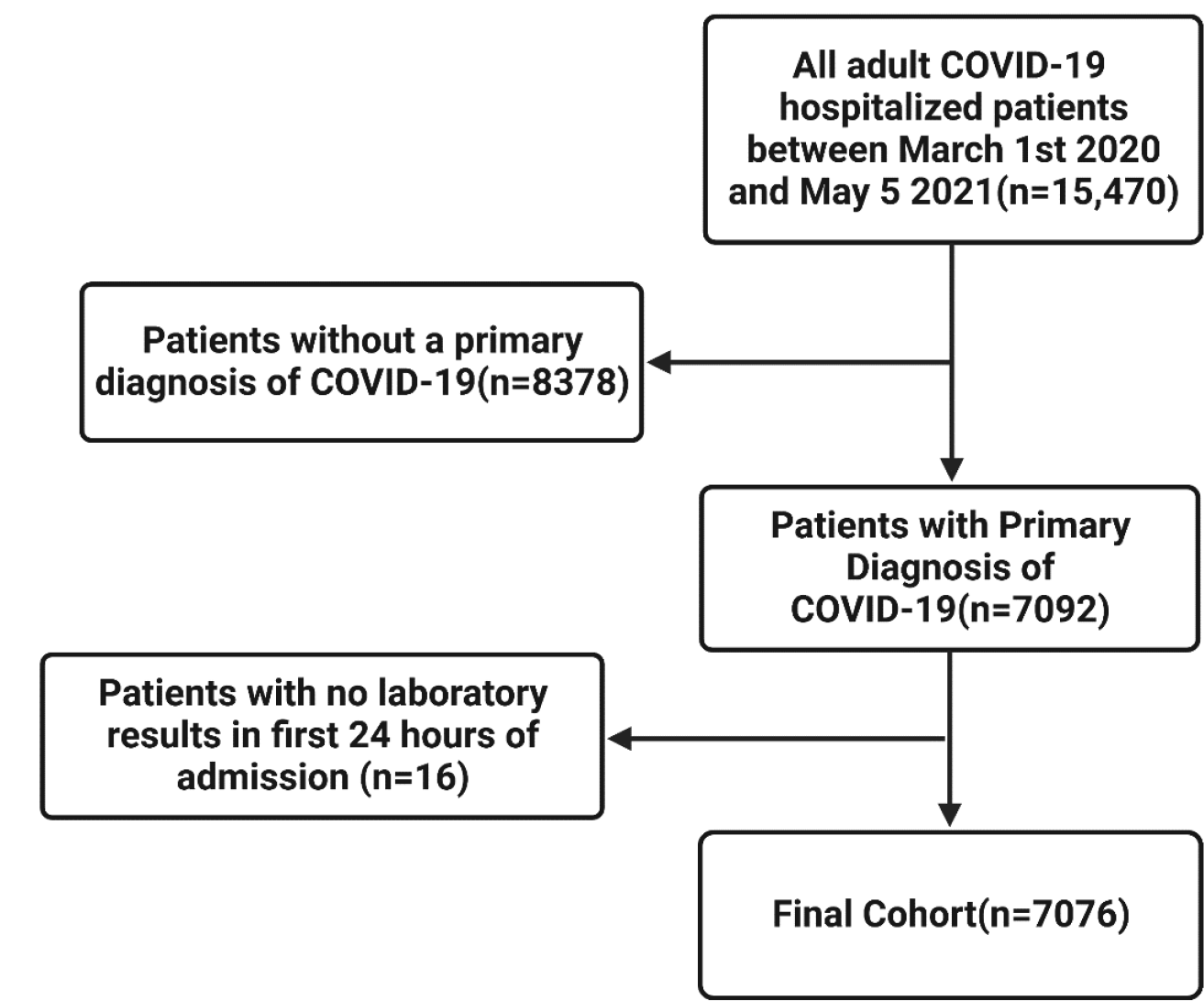
Patient selection flowchart

In the CROWN dataset database, data were labeled with an admission identification, which is specific to each patient’s hospitalization. If the patient were to be re-admitted, then they would receive a new admission identification. In this study, we treated each hospital admission as an independent sample. For patients with more than one hospitalization recorded, the mean time lapse between the discharge of a previous hospitalization and the start of the next hospitalization was 42.35 days, supporting our assumption to treat each hospitalization stay as an independent sample.

### Patient’s Data

Data for each patient contained high dimensional and longitudinal data in three main categories of interest: demographic, comorbidities, and laboratory results. Demographic data included patient age, sex, and race. Laboratory data included a periodically recorded blood panel including specific biomarkers such as IL-6, prolactin, eosinophils, and others as listed in Supplemental Table 1. Only laboratory data gathered in the first 24 hours of admission were used in the clustering analysis.

For laboratory data, we first omitted any data points that were marked as erroneous by the database. Then, we aggregated all the laboratory tests into summary statistics. We engineered features to include the minimum, maximum, mean, standard deviation, first, last, and total change of each of the laboratory tests over the first 24 hours of admission. We excluded any tests where more than 50% of the patient’s first 24h hospitalization did not have at least one value. Furthermore, for any two features with a Pearson’s correlation coefficient of greater than 90%, one feature was removed at random (Supplemental Table 1). Finally, we imputed any missing values for the remaining features using MissRanger (14), a random forest imputation package.

Primary outcomes of interest measured was mortality. Secondary outcomes of interest include myocardial infarction (MI), pulmonary embolism (PE), deep vein thrombosis (DVT), delirium, stroke, length of stay (LOS), ICU admission, and respiratory features, such as Oxygen, High-flow Oxygen, ECMO and pressor use (Applicable ICD-10 codes in Supplemental Table 2). Comorbidities to examine were chosen based on the Elixhauser Comorbidity index (15).

### Consensus Clustering

Clustering is an unsupervised machine learning tool aimed at separating heterogenous data into more homogenous subsets by grouping data according to their similarity. Simple clustering methods often assign data into groups randomly, compute a similarity index, and then aim to maximize this similarity index by rearranging patients into different groups. Then, the similarity index is recomputed with the new groups and the process is repeated. However, due to the random nature of the initial assignment, clustering algorithms are not guaranteed to reach a global optimum.

To address this issue, we used ConsensusClusterPlus (16), a consensus clustering algorithm, to group our patients into distinct subphenotypes using only the aggregated laboratory results based on the first 24 hours of their admission. Consensus clustering is an algorithm run many times, and the outcome is decided by having each run “vote” on how the clusters should be assigned. By repeatedly clustering, we significantly reduced the effect of random chance on our results. We use a partition around medoids (PAM) clustering with a Gower distance due to both simplicity and ease of interpretability. Finally, to gain further insight into the clustering, we plotted the standardized values of each feature in each cluster compared to other clusters.

### Choosing the Optimal Number of Clusters

We considered a variety of methods to determine the optimal number of clusters. Traditionally, methods such as the “elbow method” have been used to assess the optimal number of clusters. However, it has been shown that the elbow plot as well as many other traditional cluster evaluation strategies can be unreliable when used in consensus clustering algorithms (17). Instead, Șenbabaoğlu et al. propose a metric called the proportion of ambiguous clustering (PAC) to determine the optimal number of clusters (17). This method involves plotting the cumulative distribution function (CDF) curve of a consensus matrix and finding the curve that has the lowest number of ambiguous clusters. However, their study was done on low dimensional artificial data that had easy to observe clusters. When we applied the PAC method to our high dimensional data, the metric suggested that the optimal number of clusters should be the same as the number of patients (Supplemental Figure 1); that is, each patient should be its own cluster. Obviously, this would account for all heterogeneity but would not be clinically tractable. Because of this, we took this metric into account but ultimately also used clinical significance to determine the optimal number of clusters.

### Cluster Nomenclature

We used C-Reactive Protein values, which are a general indicator of the systemic inflammation level (18), to classify the clusters generated into different subphenotypes. We designated the cluster with the lowest, middle, and highest C Reactive Protein values as the hypoinflammatory cluster, intermediate cluster, and the hyperinflammatory cluster respectively.

### Standardized Mean Variable Analysis

To understand the differences in feature values between clusters identified by an unsupervised machine learning model, we computed the standardized mean differences (SMD) of each feature. The SMD is a measure of the difference in mean values between two groups, standardized by the pooled standard deviation of the groups (Supplemental Equation 1). It is useful for comparing the means of different features, as it is unitless and can be interpreted as a relative difference between the means. To compute the SMD, we first calculated the mean and standard deviation of each feature for each cluster and then used these values to standardize the feature values within each cluster. Next, we calculated the difference in standardized means between each pair of clusters and plotted these differences as SMD plots (Figure 2, Supplemental Figures 2-3). These plots provide a clear visual representation of the relative differences in feature values between the clusters and can help to identify which features are most important for distinguishing between the clusters. These results can also be used to interpret the clusters and gain insights on the underlying structure of data.

**Figure 2:**
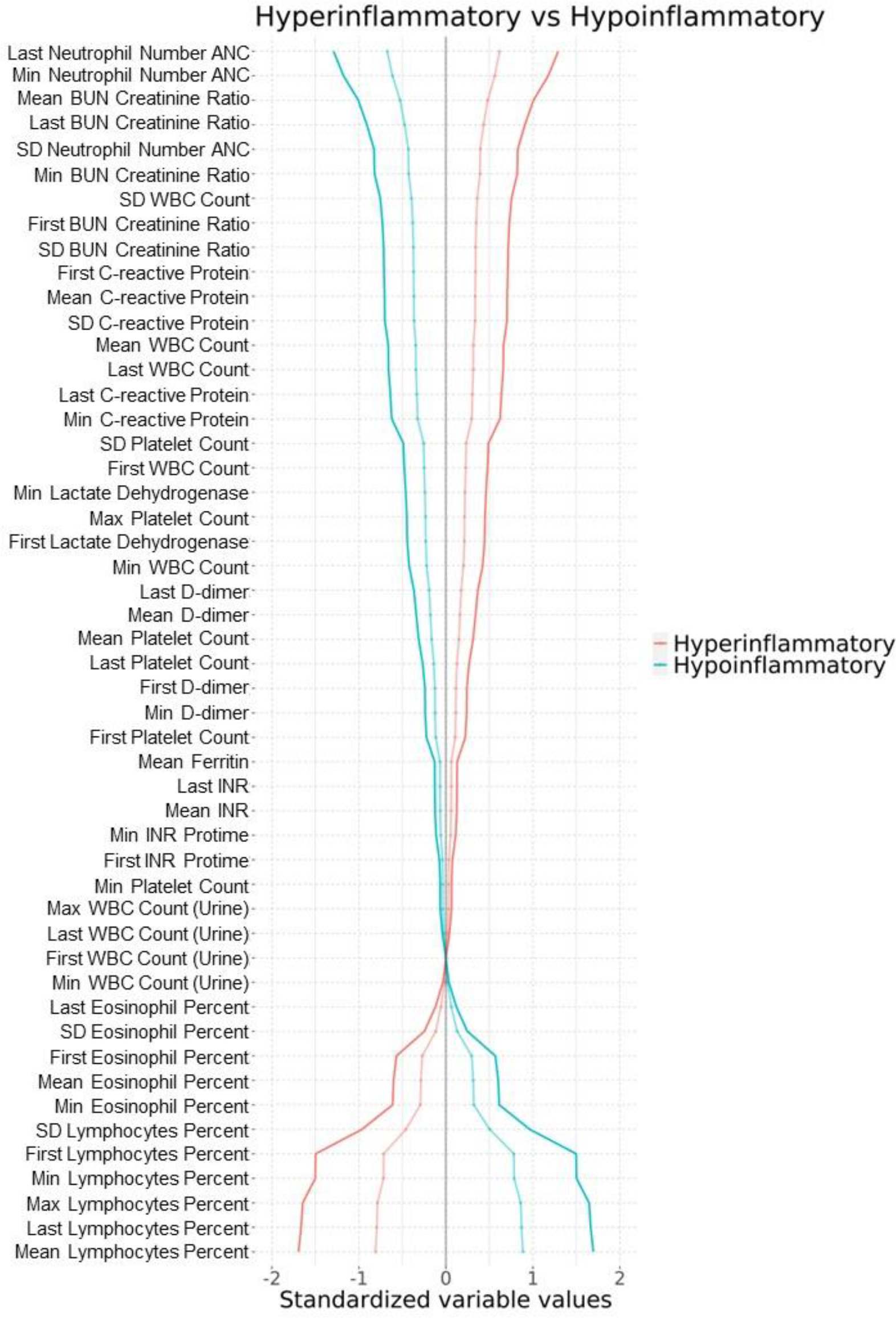
Standardized Variable Plots of Hyper vs Hypoinflammatory Clusters. Standardized mean values are computed by taking the mean of a specific feature in a group and comparing to the mean of another group (solid lines) Faded lines represent comparisons of a cluster to all patients.

### Comorbidity and Outcome Exploration

We explored how the prevalence of various outcomes differ among the clusters. Patient stay identifiers were matched with the patients from the clustering experiment to various other tables containing patient outcomes. We then counted the number of patients with the specific outcome in various cluster and conducted statistical tests to determine significance of clinical subphenotype against the outcomes of interest. We conducted Chi Squared tests for categorial variables and Mann-Whitney U tests for continuous variables. Outcomes examined are mortality, pulmonary embolism, deep vein thrombosis, myocardial infarction, and stroke. The ICD-10 codes used to identify each outcome is specified in the supplemental material (Supplemental Table 2).

Using the same method as above, we explored the association of phenotype assignment with preexisting comorbidities. We chose to examine comorbidities that are listed in the Elixhauser Comorbidity Index.

## Results

Starting with 15,470 adult COVID-19 positive patients, we followed the patient selection criteria as described above, which resulted in 7076 patients being included in the study (Figure 1). Following the cluster procedure above, we generated hypoinflammatory, intermediate, and hyperinflammatory subphenotypes with 2090, 3076, and 1910 patients in each subphenotype respectively. The hypoinflammatory patients had, on average, a younger mean age, a lower number of total comorbidities, lower rates of ICU admission, and a lower average hospitalization stay (Table 1).

**Table 1:** Cluster descriptions

| Results | Hyperinflammatory | Intermediate | Hypoinflammatory | p-value |
| --- | --- | --- | --- | --- |
| Cluster Descriptions |  |  |  |  |
| Cluster Size | 2090 | 3076 | 1910 | 4.97E-08 |
| # Of ICU Admits | 250 (12.0%) | 180 (5.9%) | 50 (2.6%) | 2.86E-32 |
| # Non Male | 915 (43.8%) | 1497 (48.7%) | 1066 (55.8%) | 4.53E-13 |
| median LOS(days) | 7 | 5 | 4 | 1.03E-53 |
| survival | 1562 (74.7%) | 2904 (94.4%) | 1865 (97.6%) | 9.17E-127 |
| Age | 67 (3.2%) | 60 (2.0%) | 54 (2.8%) | 4.03E-107 |
| Respiratory Features |  |  |  |  |
| Ventilator | 445 (21.3%) | 297 (9.7%) | 71 (3.7%) | 1.87E-70 |
| Oxygen | 1366 (65.4%) | 1876 (61.0%) | 743 (38.9%) | 7.14E-73 |
| ECMO | 15 (0.7%) | 7 (0.2%) | 3 (0.2%) | 0.00030055 |
| high flow | 560 (26.8%) | 458 (14.9%) | 89 (4.7%) | 1.09E-81 |
| pressors | 368 (17.6%) | 265 (8.6%) | 62 (3.2%) | 3.92E-53 |
| Comorbidities |  |  |  |  |
| Depression | 577 (27.6%) | 842 (27.4%) | 658 (34.6%) | 4.97E-08 |
| Anemia Deficiency | 1126 (53.9%) | 1503 (48.9%) | 986 (51.8%) | 0.00167542 |
| Hypertension | 1557 (74.5%) | 2171 (70.6%) | 1250 (65.7%) | 7.06E-09 |
| Weightloss | 511 (24.4%) | 528 (17.2%) | 364 (19.1%) | 6.70E-10 |
| lymphoma | 55 (2.6%) | 84 (2.7%) | 60 (3.2%) | 0.57119295 |
| Coagulation deficiency | 510 (24.4%) | 737 (2.4%) | 459 (24.1%) | 0.93944303 |
| Alcohol use disorder | 115 (5.5%) | 214 (.7%) | 236 (12.4%) | 2.06E-16 |
| Congestive Heart Failure | 659 (31.5%) | 780 (25.4%) | 451 (23.7%) | 1.20E-08 |
| Renal failure | 723 (34.6%) | 916 (29.8%) | 517 (27.2%) | 1.18E-06 |
| Perivascular Disease | 470 (22.5%) | 557 (18.1%) | 357 (18.8%) | 0.00030055 |
| Solid Tumor without Metastasis | 359 (17..2%) | 436 (14.2%) | 211 (11.1%) | 2.59E-07 |
| AIDS | 25 (1.2%) | 73 (2.4%) | 52 (2.7%) | 0.00152586 |
| Paralysis | 240 (11.5%) | 243 (7.9%) | 165 (8.7%) | 4.73E-05 |
| Pulmonary circulation disorders | 250 (1.2%) | 305 (9.9%) | 225 (11.8%) | 0.03184051 |
| Hypertension with chronic complications | 973 (46.6%) | 1211 (39.4%) | 703 (36.9%) | 4.59E-10 |
| Chronic Peptide Ulcer | 103 (4.9%) | 135 (4.4%) | 95 (0.5%) | 0.5361094 |
| Psychoses | 174 (8.3%) | 292 (9.5%) | 270 (14.2%) | 9.79E-10 |
| Obesity | 771 (36.9%) | 1330 (43.3%) | 863 (45.3%) | 6.58E-08 |
| Blood loss anemia | 158 (7.6%) | 241 (7.8%) | 162 (8.5%) | 0.52275007 |
| Chronic pulmonary disease | 697 (33.3%) | 900 (29.3%) | 622 (32.7%) | 0.00312582 |
| Drug abuse | 128 (6.1%) | 203 (6.6%) | 272 (14.3%) | 8.10E-25 |
| Hypothyroidism | 404 (19.3%) | 491 (1.6%) | 276 (14.5%) | 0.00010953 |
| Metastatic Cancer | 224 (10.7%) | 268 (8.7%) | 146 (7.7%) | 0.00258804 |
| Fluid electrolyte disorders | 1656 (79.2%) | 2155 (70.1%) | 1284 (67.4%) | 5.38E-18 |
| Liver Disease | 278 (13.3%) | 490 (15.9%) | 390 (20.5%) | 4.84E-09 |
| Rheumatoid arthritis | 154 (7.4%) | 192 (6.2%) | 132 (6.9%) | 0.27180647 |
| Neurological disorders | 699 (33.4%) | 881 (28.7%) | 595 (31.3%) | 0.00108518 |
| Diabetes with Chronic complications | 834 (39.9%) | 1114 (36.2%) | 645 (33.9%) | 0.00032622 |
| Valvular Disease | 341 (16.3%) | 427 (13.9%) | 262 (13.8%) | 0.02653807 |
| Diabetes without chronic complications | 839 (40.1%) | 1266 (41.2%) | 752 (39.5%) | 0.47546544 |

To examine the features that drove the cluster decision algorithm, we plotted standardized variable values comparing the three clusters (Figure 2, Supplemental Figure 2 and Supplemental Figure 3). The hypoinflammatory clusters are characterized by high lymphocyte values but low neutrophil values, whereas the hyperinflammatory values are characterized by low lymphocyte values but high neutrophil values. Upon examining the levels of biomarkers, we discovered that many biomarkers have significant differences across the clusters (Table 2).

**Table 2:** Results and Outcomes by cluster

|  | Hyperinflammatory | Intermediate | Hypoinflammatory | p-value |
| --- | --- | --- | --- | --- |
| Cluster Size | 2090 | 3076 | 1910 |  |
| MI | 226 (10.8%) | 212 (6.89%) | 98 (5.13%) | 1.652e-11 |
| PE | 125 (5.98%) | 119 (3.87%) | 97 (5.08%) | 0.001942 |
| DVT | 135 (6.45%) | 117 (3.80%) | 84 (4.40%) | 4.285e-05 |
| Delirium | 226 (10.8%) | 212 (6.89%) | 98 (5.13%) | 1.652e-11 |
| Stroke | 63 (3.01%) | 62 (2.02%) | 32 (1.68%) | 0.009648 |

To further define the clusters, we examined a detailed breakdown of the clinical outcomes during hospitalization in each cluster (Table 3). We found that the hyperinflammatory class had worse outcomes than the intermediate cluster, which, in turn, had worse outcomes than the hypoinflammatory subphenotypes. Of note, this relationship was not noted for pulmonary embolism.

**Table 3:**
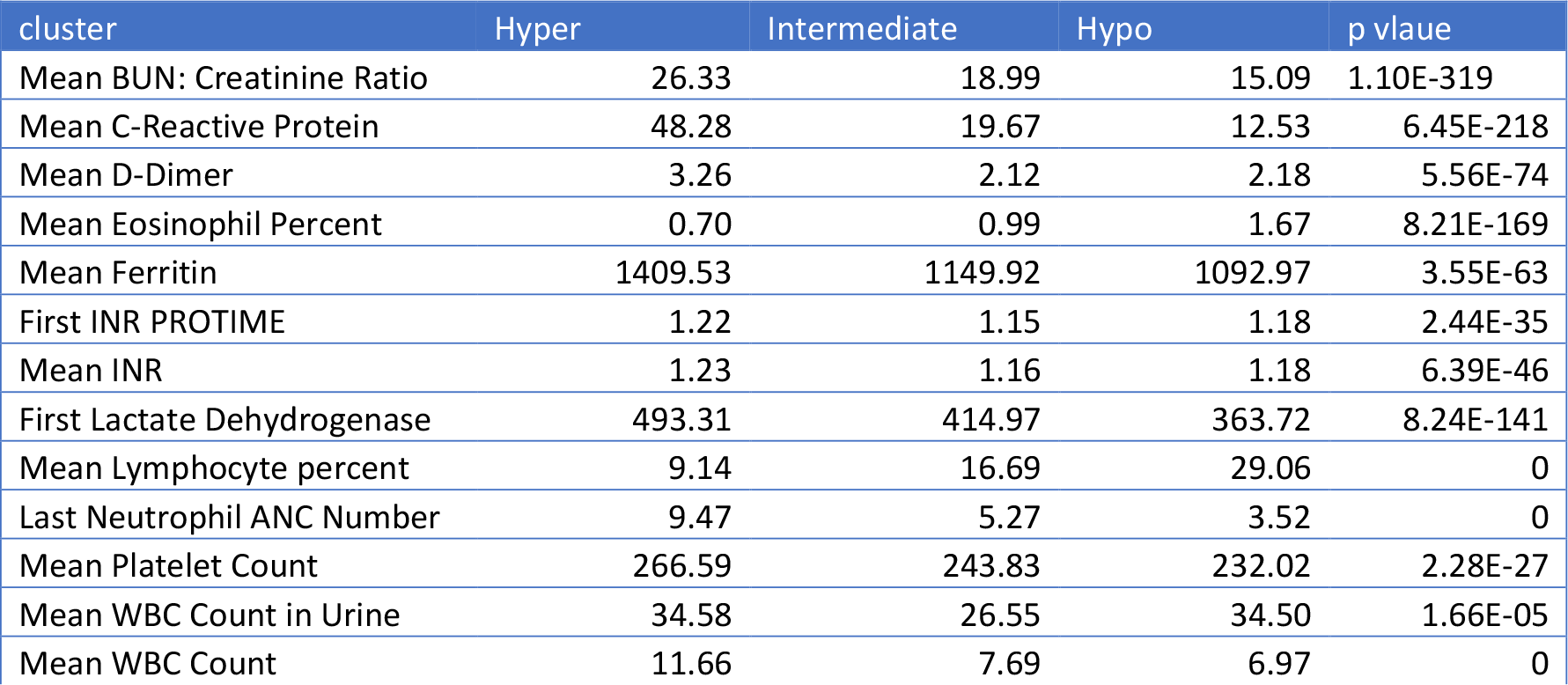
Mean of biomarker features used in clustering

## Discussion

In this study, we clustered COVID-positive patients into three distinct subphenotypes based on inflammatory biomarkers collected within the first 24 hours of admittance to hospital. We found significant correlations between inflammatory subphenoypes and the occurrence of key complications during hospitalization (stroke, myocardial infarctions (MI), deep-vein thrombosis (DVT), delirium, and mortality).

The hypoinflammatory subphenotypes are characterized by higher levels of lymphocytes and eosinophils despite having a lower C-reactive protein levels, which characterize overall inflammation. The hyperinflammatory clusters have the highest BUN (creatinine) ration as well as the highest neutrophil counts (Figure 2).

### Physiology of disease

In characterizing the features associated with the discrete clusters, we identified features that appear consistent with current understandings of COVID-19 pathophysiology. We noted relative eosinophilia associated with the hypoinflammatory subphenotype. Notably, eosinopenia has been associated with poor outcomes in COVID-19 and recovery of eosinophil counts has been putatively associated with a favorable recovery trajectory (19). Moreover, we observed the hyperinflammatory group had higher BUN-Creatine ratios. A common clinical measure of renal function, increases in BUN and serum creatinine in patients with COVID-19 have been strongly associated with increased rates of adverse outcomes (20). The causal relationship between renal function, volume status, and COVID-19 remains to be elucidated, but it does appear this may be a hallmark of worse relative outcome.

### Clinical Utility

Early recognition of COVID-19 inflammatory subphenotypes may facilitate clinical prognostication, enable better clinical decision-making, and manifest in better patient outcomes. By having only three subphenotypes that can be easily distinguished by biomarkers, the subphenotypes identified by our study may be easier to recognize than existing subphenotypes. For instance, Dubowski et al. produced 6 subphenotypes based on over 30 biomarkers as well as vital signs features (7). This is a significantly more complicated scheme than our subphenotypes, making it harder for providers to determine which subphenotype a given patient would belong in. On the contrary, a provider can easily determine whether a given patient would belong in our hyperinflammatory subphenotype by obtaining a blood panel and examining a few key features.

In addition, our results suggest the possibility of a more personalized care routine for dexamethasone use. Severe COVID-19 patients may experience inflammatory organ injury due to strong host inflammatory responses, and dexamethasone is believed to mitigate this damage by attenuating the host inflammatory response (2,21,22). At the time of writing, current guidelines are based on the RECOVERY trial, which showed dexamethasone improved outcomes for hospitalized adult patients if they require oxygen therapy (23,24). However, literature has shown that dexamethasone can lead to a variety of potential side effects, such as dexamethasone-induced hypertension (25,26), osteonecrosis of the femoral head (27,28), ventricular hypertrophy (29), diabetes (30), and other complications. Our results show that although the 38.9% of hypoinflammatory patients receive oxygen therapy, these patients experience a different type of inflammatory response than the hyperinflammatory patients. Taken in conjunction with their better outcomes, it questions the current one-size-fits-all approach to glucocorticoids. More importantly, these results also suggest that any patient in the hyperinflammatory subphenotype may benefit from glucocorticoid treatments, even if they are not currently on oxygen.

### Clinical Trial Implications

Our results carry significant impact for future design and implementation of anti-inflammatory and immune modulating clinical trials for COVID -19 patients. Even though these medications carry significant adverse effects (31), they continue to be prescribed for many hospitalized COVID-19 patients (32). Our results point to a personalized approach that could be used to select patients in clinical trials of COVID-19 therapies. The inflammatory subphenotypes suggest that anti-inflammatory and immune modulating therapies might be highly beneficial when treating patient in the hyperinflammatory subphenotype but ineffective or detrimental if used in patients in the hypoinflammatory subphenotype. Future studies are needed to verify this hypothesis.

### Strengths

The COVID-CROWN database by the Precision Medicine Analytics Platform offers highly detailed information regarding the patients all throughout their hospital stay. The dataset contained data from 7076 study eligible patients who were treated in 5 different hospitals, which increases the generalizability of the results compared to single hospital studies. This highly detailed data allowed us to conduct an in-depth exploration of how the patient’s inflammatory status was related to their disease trajectory throughout their inpatient stay. We demonstrated that a patient’s outcome was tangibly related to their inflammation status and that the hyperinflammatory subgroups had significantly worse disease trajectories. In addition, our subphenotypes are easily identifiable using a few key biomarker characteristics, making them potentially suitable to guide clinical decision making.

Unlike other subphenotyping exercises, our study examined patients spanning over a year, which contained several waves of COVID involving different strains of SARS-CoV-2. This suggests that our study can be more resistant to bias for one strain. However, additional work should be done to update our model in light of more recent strains.

### Limitations

We acknowledge that our study has limitations. One potential limitation is the lack of external validation. We were unable to overcome this due to challenges in identifying large-scale open access databases that contain similar detailed information as in the COVID-CROWN database. We also acknowledge that the COVID-19 pandemic response moved at a very fast pace. Certain treatments that may not have been available at the beginning of the pandemic became widely used towards the later stages. These treatments were not considered in our modeling approach. Additionally, there are limitations towards picking the number of optimal clusters. In our methodology section, we present evidence as to why traditional methods such as elbow plots or silhouette scores are inappropriate for consensus clustering. Furthermore, scores based on sampling or bootstrapping the data (i.e. Jaccard Coefficient) are also inappropriate due to the repeated sampling procedure already in consensus clustering.

## Conclusion

In conclusion, we find three subphenotypes of COVID-19 that are easily distinguishable within the first 24 hours of hospital admission. The hyperinflammatory subphenotypes have significant increases when adverse outcomes occurred, including mortality, pulmonary embolism, stroke, deep vein thrombosis, delirium and myocardial infarction. Additional studies are needed to confirm the external validity of these results.

## Data Availability

The data used were part of JH-CROWN: The COVID Precision Medicine Analytics Platform Registry.Data and analysis code will be made available upon researcher request as allowable by the terms of the registry and the Johns Hopkins Medicine Institutional Review Board (IRB)

