## Supplemental Information for "Unsupervised Machine Learning Unveil Easily Identifiable Subphenotypes of COVID-19 With Differing Disease Trajectories"

### Supplemental figures

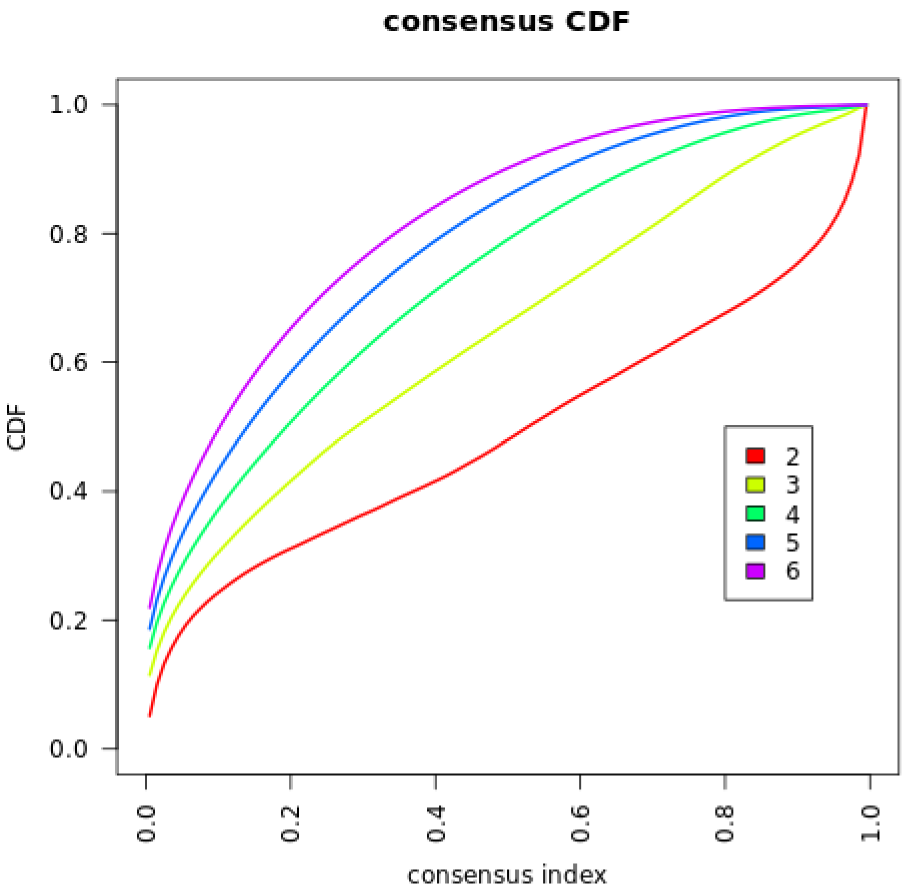

*Supplemental Figure 1: Cumulative Distribution Function plotted against consensus index. The Consensus index represents how often two samples are placed together in the same cluster. A consensus value of one indicates that the two samples are always grouped together while a value of 0 indicates that two samples are never grouped together. Percent of Ambiguous Clusters (PAC) is a measure of how much of the CDF falls between a consensus index between 0.1 and 0.9. The plot above shows that as we increase the number of clusters, it's clear that the PAC will always decrease. Hence choosing the minimal PAC will lead to a cluster size equal to the number of samples.*

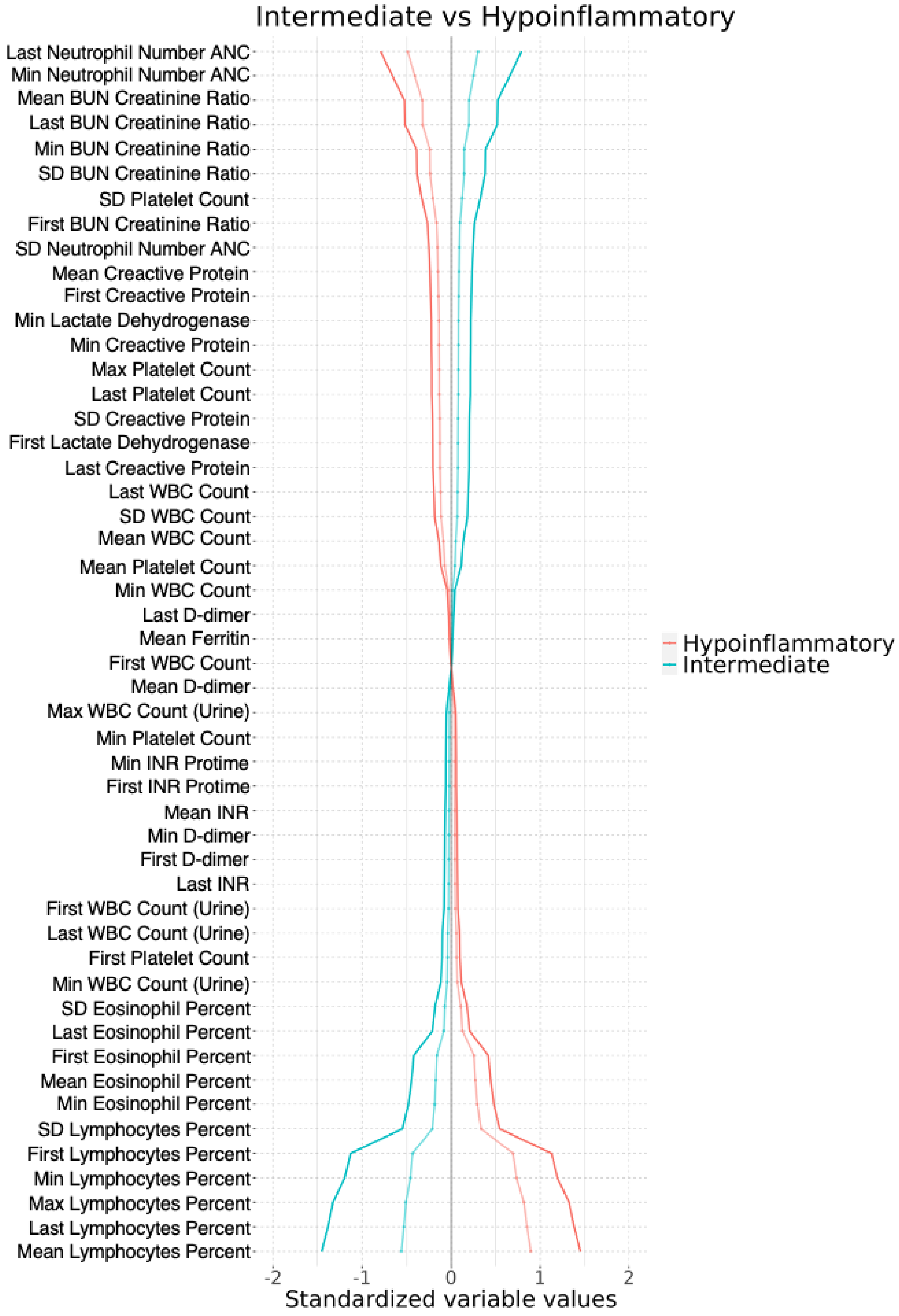
Intermediate vs Hypoinflammatory
Last Neutrophil Number ANC
Min Neutrophil Number ANC
Mean BUN Creatinine Ratio
Last BUN Creatinine Ratio
Min BUN Creatinine Ratio
SD BUN Creatinine Ratio
SD Platelet Count
First BUN Creatinine Ratio
SD Neutrophil Number ANC
Mean Creactive Protein
First Creactive Protein
Min Lactate Dehydrogenase
Min Creactive Protein
Max Platelet Count
Last Platelet Count
SD Creactive Protein
First Lactate Dehydrogenase
Last Creactive Protein
Last WBC Count
SD WBC Count
Mean WBC Count
Mean Platelet Count
Min WBC Count
Last D-dimer
Mean Ferritin
First WBC Count
Mean D-dimer
Max WBC Count (Urine)
Min Platelet Count
Min INR Protime
First INR Protime
Mean INR
Min D-dimer
First D-dimer
Last INR
First WBC Count (Urine)
Last WBC Count (Urine)
First Platelet Count
Min WBC Count (Urine)
SD Eosinophil Percent
Last Eosinophil Percent
First Eosinophil Percent
Mean Eosinophil Percent
Min Eosinophil Percent
SD Lymphocytes Percent
First Lymphocytes Percent
Min Lymphocytes Percent
Max Lymphocytes Percent
Last Lymphocytes Percent
Mean Lymphocytes Percent
-2
-1
0
1
2
Standardized variable values
Hypoinflammatory
Intermediate

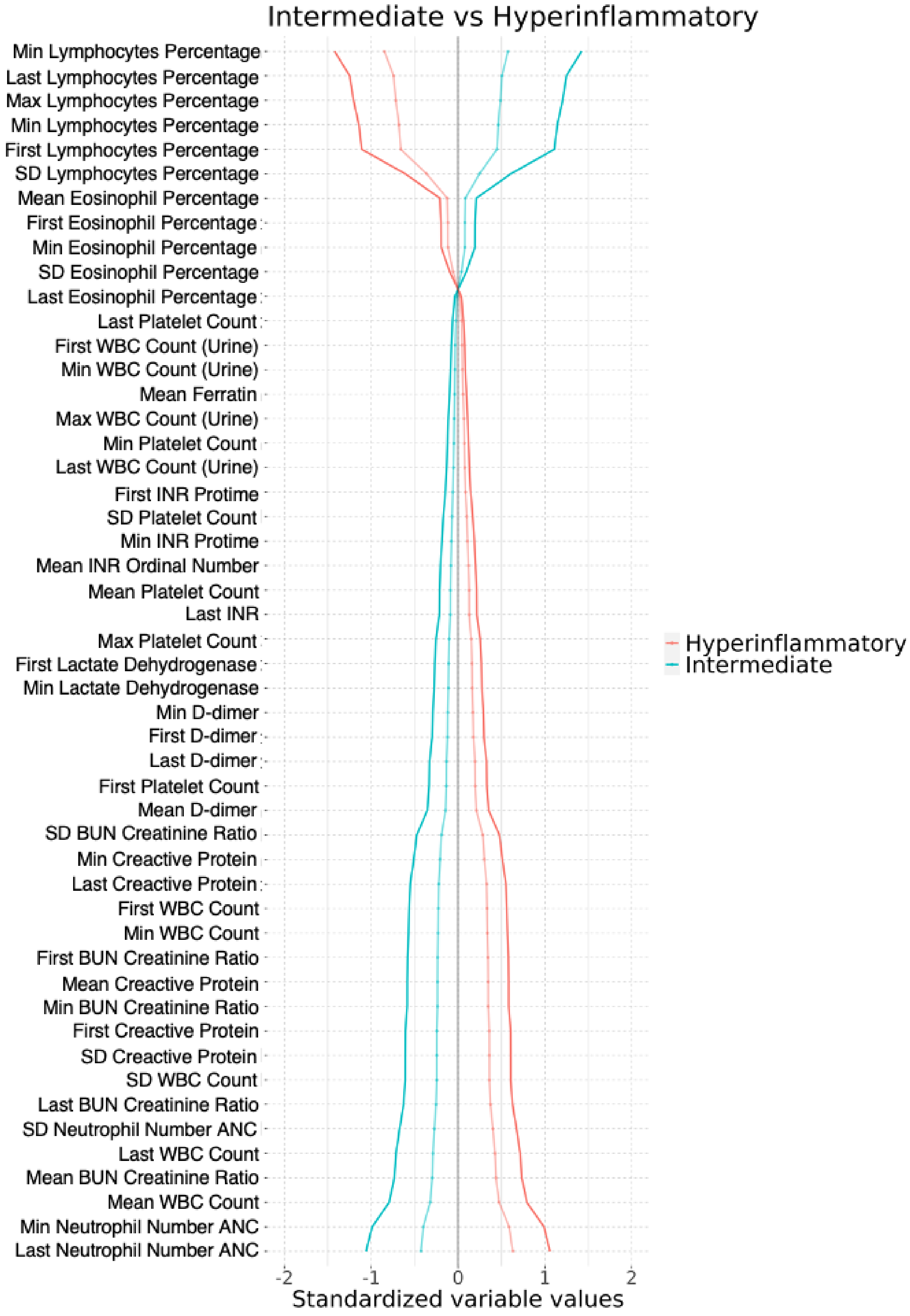
Intermediate vs Hyperinflammatory
Min Lymphocytes Percentage
Last Lymphocytes Percentage
Max Lymphocytes Percentage
Min Lymphocytes Percentage
First Lymphocytes Percentage
SD Lymphocytes Percentage
Mean Eosinophil Percentage
First Eosinophil Percentage
Min Eosinophil Percentage
SD Eosinophil Percentage
Last Eosinophil Percentage
Last Platelet Count
First WBC Count (Urine)
Min WBC Count (Urine)
Mean Ferratin
Max WBC Count (Urine)
Min Platelet Count
Last WBC Count (Urine)
First INR Protime
SD Platelet Count
Min INR Protime
Mean INR Ordinal Number
Mean Platelet Count
Last INR
Max Platelet Count
First Lactate Dehydrogenase
Min Lactate Dehydrogenase
Min D-dimer
First D-dimer
Last D-dimer
First Platelet Count
Mean D-dimer
SD BUN Creatinine Ratio
Min Creactive Protein
Last Creactive Protein
First WBC Count
Min WBC Count
First BUN Creatinine Ratio
Mean Creactive Protein
Min BUN Creatinine Ratio
First Creactive Protein
SD Creactive Protein
SD WBC Count
Last BUN Creatinine Ratio
SD Neutrophil Number ANC
Last WBC Count
Mean BUN Creatinine Ratio
Mean WBC Count
Min Neutrophil Number ANC
Last Neutrophil Number ANC
-2
-1
0
1
2
Standardized variable values
Hyperinflammatory
Intermediate

### Supplemental Tables

| Column1 | Mean | Standard Deviation | First | Last | Min | Max |
|---|---|---|---|---|---|---|
| CREACTIVE_PROTEIN | CREACTIVE_PROTEIN_Mean | CREACTIVE_PROTEIN_sd | CREACTIVE_PROTEIN_First | CREACTIVE_PROTEIN_Last | CREACTIVE_PROTEIN_Min | |
| DDIMER | DDIMER_Mean | | DDIMER_First | DDIMER_Last | DDIMER_Min | |
| FERRITIN | FERRITIN_Mean | | | | | |
| LACTATE_DEHYDROGENASE | LACTATE_DEHYDROGENASE_Mean | | LACTATE_DEHYDROGENASE_First | | | |
| LYMPHOCYTES_percent | LYMPHOCYTES_percent_Mean | LYMPHOCYTES_percent_sd | LYMPHOCYTES_percent_First | LYMPHOCYTES_percent_Last | LYMPHOCYTES_percent_Min | LYMPHOCYTES_percent_Max |
| PLATELET_COUNT | PLATELET_COUNT_Mean | PLATELET_COUNT_sd | PLATELET_COUNT_First | PLATELET_COUNT_Last | PLATELET_COUNT_Min | PLATELET_COUNT_Max |
| NEUTROPHIL_NUMBER_ANC | | NEUTROPHIL_NUMBER_ANC_sd | | NEUTROPHIL_NUMBER_ANC_Last | NEUTROPHIL_NUMBER_ANC_Min | NEUTROPHIL_NUMBER_ANC_Max |
| EOSINOPHIL_percent | EOSINOPHIL_percent_Mean | EOSINOPHIL_percent_sd | EOSINOPHIL_percent_First | EOSINOPHIL_percent_Last | EOSINOPHIL_percent_Min | |
| BUN_._CREATININE_RATIO | BUN_._CREATININE_RATIO_Mean | BUN_._CREATININE_RATIO_sd | BUN_._CREATININE_RATIO_First | BUN_._CREATININE_RATIO_Last | BUN_._CREATININE_RATIO_Min | |
| INR._PROTIME | | | INR._PROTIME_First | | INR._PROTIME_Min | |
| WBC_COUNT_URINE | | | WBC_COUNT_URINE_First | WBC_COUNT_URINE_Last | WBC_COUNT_URINE_Min | WBC_COUNT_URINE_Max |
| WBC_ord_num_value | WBC_ord_num_value_Mean | WBC_ord_num_value_sd | WBC_ord_num_value_First | WBC_ord_num_value_Last | WBC_ord_num_value_Min | |
| INR_ord_num_value | INR_ord_num_value_Mean | | | INR_ord_num_value_Last | | |

*Supplemental Table 1: Describes which features were selected and kept in feature selection*

*Supplemental Table 2:ICD 10 codes used to explore outcomes*

| Condition | ICD-10 Code | Count | Total Count | Unique Total Count |
|---|---|---|---|---|
| DVT | I82 | 444 | 444 | 336 |
| PE | I26 | 287 | 354 | 341 |
| | T81.7 | 3 | | |
| | T82.8 | 49 | | |
| | T79.0 | 2 | | |
| | T79.1 | 1 | | |
| | T80.0 | 0 | | |
| MI | I21 | 442 | 548 | 536 |
| | I22 | 1 | | |
| | I23 | 0 | | |
| | I24 | 97 | | |

| | | |
|---|---|---|
| Stroke: | I60<br>I61<br>I63<br>I65<br>I66<br>G45 | 18<br>45<br>103<br>33<br>3<br>15 |

Supplemental Table 3. Race Results

| cluster | first_race | n |
|---|---|---|
| 1 | Am Indian | 6 |
| 1 | Asian | 160 |
| 1 | Black | 1128 |
| 1 | NotDisclose | 1 |
| 1 | Other | 615 |
| 1 | Other Pacifi | 1 |
| 1 | Pac Islander | 1 |
| 1 | Unknown | 13 |
| 1 | White | 1148 |
| 1 | NA | 3 |
| 2 | Am Indian | 4 |
| 2 | Asian | 128 |
| 2 | Black | 598 |
| 2 | Hispanic | 1 |
| 2 | NotDisclose | 2 |
| 2 | Other | 335 |
| 2 | Other Pacifi | 1 |
| 2 | Pac Islander | 2 |
| 2 | Unknown | 15 |
| 2 | White | 1004 |
| 3 | Am Indian | 3 |
| 3 | Asian | 65 |
| 3 | Black | 859 |
| 3 | Hispanic | 1 |
| 3 | NotDisclose | 3 |
| 3 | Other | 370 |
| 3 | Pac | 1 |

| | | |
|---|---|---|
| | Islander | |
| 3 | Unknown | 7 |
| 3 | White | 593 |
| 3 | NA | 8 |

### Supplemental Equation

Standardized Mean Difference

$$SMD = \frac{\overline{X_1} - \overline{X_2}}{\sqrt{(S_1^2 + S_2^2)/2}}$$

Where $\overline{X_1}$ and $\overline{X_2}$ represent the mean of a variable of interest in group 1 and two respectively while $S_1$ and $S_2$ represent the variance.
